# Adaptive Frequency-Spatial Dual-Stream Network (AFS-DSN) for Nasal and Paranasal Sinus CT Segmentation

**DOI:** 10.64898/2026.04.19.26351206

**Authors:** Shu-Yen Wan, Wen-Yu Chen

## Abstract

Accurate segmentation of nasal and paranasal sinus structures from CT scans is critical for surgical planning and treatment evaluation in rhinology. However, the complex anatomical topology and thin-wall boundaries of these structures pose significant challenges for automated segmentation methods. We propose AFS-DSN (Adaptive Frequency-Spatial Dual-Stream Network), a novel deep learning architecture that integrates multi-scale wavelet decomposition with spatial feature learning for binary segmentation of the nasal cavity complex. Our method employs a dual-stream encoder with frequency branch utilizing three wavelet scales (db1, db2, db4) to capture 24 frequency sub-bands, enabling enhanced boundary detection in anatomically challenging regions. Cross-domain attention and adaptive routing mechanisms dynamically fuse spatial and frequency features based on local tissue characteristics. We formulate the task as binary segmentation where all five anatomical structures (maxillary sinus, sphenoid sinus, ethmoid sinus, frontal sinus, and nasal cavity) are treated as a unified foreground region against the background, prioritizing clinical boundary detection over individual structure differentiation. Evaluated on the NasalSeg dataset (130 CT volumes) with a 70/15/15 train/validation/test split, AFS-DSN achieves 94.34% ± 2.30% overall Dice coefficient with statistically significant improvements in thin-wall regions (91.34% vs. 90.57% baseline, *p*=0.004) and statistically significant improvement in Surface Dice at 1mm tolerance (0.874 vs. 0.868 baseline, *p*=0.010), demonstrating enhanced boundary precision while maintaining sub-second inference time, making the method suitable for surgical planning applications where sub-millimeter accuracy is clinically relevant. To address concerns regarding model complexity, we further introduce AFS-DSN-Lite, a parameter-efficient variant (27.41M parameters) that achieves comparable performance (94.37% Dice) through depthwise separable convolutions, and validate robustness via 3-fold cross-validation (mean Dice: 94.59% ± 0.31%).

## 1 Introduction

Chronic rhinosinusitis and other sinonasal diseases are highly prevalent and often lead to significant impairment in patients’ quality of life. Computed tomography (CT) is routinely used for diagnosis, surgical planning, and postoperative follow-up, as it provides detailed visualization of the nasal cavity and paranasal sinuses. However, manual delineation of sinonasal structures on 3D CT volumes is labor-intensive, time-consuming, and subject to inter-observer variability, which limits its practicality in large-scale clinical workflows and quantitative analysis. Automatic, accurate, and robust segmentation of the nasal cavity and paranasal sinuses is therefore a critical prerequisite for computer-assisted diagnosis, objective disease scoring, and image-guided endoscopic sinus surgery.

Despite recent advances in deep learning for medical image segmentation, automatic sinonasal segmentation remains challenging. The nasal cavity and paranasal sinuses exhibit complex and highly variable anatomy, with thin bony walls, narrow air channels, and frequent postoperative deformities. CT images often suffer from low soft-tissue contrast, partial volume effects, and streak artifacts from dental fillings or implants, which can blur mucosal boundaries and obscure small structures. Conventional 3D convolutional neural networks (CNNs), such as U-Net and its variants, primarily learn spatial-domain features and tend to over-smooth fine details, leading to boundary leakage or under-segmentation in regions with thin structures or subtle intensity transitions.

More recently, transformer-based architectures and state-space models have been introduced to capture long-range dependencies and global context in medical images. Methods such as UN-ETR [9] and Swin-UNETR [10] combine CNN-like encoders with vision transformers to enlarge the receptive field, while Mamba-based networks such as U-Mamba [13] and SegMamba [14] leverage sequence modeling to improve feature integration. Although these approaches achieve strong performance on various organ and lesion segmentation tasks, they still operate predominantly in the spatial domain and do not explicitly model frequency-domain information.

The frequency domain provides complementary information to spatial representations: while spatial features capture local intensity patterns and textures, frequency features explicitly encode edge sharpness, structural periodicity, and multi-scale anatomical details through wavelet decomposition. For sinonasal CT segmentation, frequency-domain processing is particularly beneficial because: (1) thin bony walls and mucosal boundaries manifest as high-frequency components that can be enhanced through targeted wavelet filtering; (2) low-frequency sub-bands can suppress noise and artifacts while preserving global anatomical structure; and (3) multi-scale wavelet analysis naturally captures the hierarchical organization of sinonasal anatomy from large sinuses to fine ostia. Despite these theoretical advantages, existing spatial–frequency networks are mostly designed for 2D images or generic anatomies and have not been tailored to the unique challenges of 3D sinonasal CT segmentation.

To address these limitations, this paper proposes AFS-DSN, an Adaptive Frequency–Spatial Dual-Stream Network for automatic segmentation of the nasal cavity and paranasal sinuses from 3D CT volumes. Building upon a 3D U-Net–style spatial encoder–decoder backbone, AFS-DSN introduces a parallel frequency branch based on multi-scale 3D wavelet decomposition, a cross-domain attention module for bidirectional interaction between spatial and frequency streams, and an adaptive router that dynamically fuses the two domains according to local anatomical complexity. The network is trained and evaluated on the NasalSeg dataset [15], the first large-scale, publicly available sinonasal CT dataset with voxel-wise annotations of five anatomical structures.

The main contributions of this work are threefold:

- A novel adaptive frequency–spatial dual-stream architecture (AFS-DSN) is proposed for 3D nasal cavity and paranasal sinus segmentation, explicitly modeling complementary spatial and frequency-domain representations via multi-scale wavelet analysis.
- A cross-domain attention module and content-aware adaptive router are designed to enable bidirectional feature exchange and dynamic fusion between spatial and frequency streams, leading to improved boundary preservation and robustness in challenging sinonasal regions.
- Extensive experiments on the NasalSeg dataset demonstrate that AFS-DSN achieves promising performance, with a Dice score of 94.34% ± 2.30% and HD95 of 1.189 mm on the independent test set, significantly outperforming strong baselines including nnU-Net (+0.79%, *p <*0.05), UNETR, Swin-UNETR, U-Mamba, and SegMamba. We further introduce AFS-DSN-Lite (27.41M parameters), a parameter-efficient variant achieving 94.37% Dice through depthwise separable convolutions, and validate robustness via 3-fold cross-validation (94.59% ± 0.31%).

## 2 Related Work

This section examines key developments across diverse fields relevant to automated sinonasal segmentation, including clinical diagnostic challenges, the evolution of deep learning-based segmentation architectures, and recent advances in multi-stream and frequency-domain medical image analysis.

### 2.1 Clinical Background: Nasal and Paranasal Sinus Anatomy

Chronic rhinosinusitis (CRS) is a prevalent inflammatory condition affecting the nasal cavity and paranasal sinuses, impacting approximately 10% of the global population. Accurate delineation of sinonasal anatomical structures from computed tomography (CT) scans is critical for surgical planning, particularly for functional endoscopic sinus surgery (FESS). However, manual segmentation remains time-consuming and subject to substantial inter-observer variability, especially for small structures such as the inferior turbinate, middle meatus, and ethmoidal air cells, where anatomical boundaries are often ill-defined or blurred in CT images.

Empty Nose Syndrome (ENS) represents a debilitating post-surgical complication characterized by paradoxical nasal obstruction despite anatomically patent airways [18]. Clinical studies have demonstrated its substantial impact on patients’ quality of life, including associations with psychiatric symptomatology and sleep disturbances [20]. Early detection of at-risk anatomical patterns requires precise quantification of residual turbinate volume and nasal airway geometry, which demands automated, high-fidelity segmentation algorithms capable of handling anatomical variability and image quality variations across patient populations.

Recent deep learning approaches have been applied specifically to sinonasal CT segmentation. Massey et al. [11] validated a CNN-based algorithm for automated 3D paranasal sinus segmentation in chronic rhinosinusitis patients, demonstrating strong correlation with clinical scoring systems. Whangbo et al. [16] compared multiple 3D U-Net variants for multi-class paranasal sinus segmentation, highlighting the challenge of delineating individual sinus structures. Despite these advances, prior methods have not explicitly modeled frequency-domain information, which is particularly relevant for the thin bony boundaries characteristic of sinonasal anatomy.

### 2.2 Deep Learning for Medical Image Segmentation

The advent of fully convolutional networks (FCN) [1] marked a paradigm shift in dense prediction tasks, enabling end-to-end learning of pixel-wise semantic labels directly from image data. U-Net [2], introduced for biomedical image segmentation, combined a contracting encoder path with an expansive decoder path featuring skip connections to recover spatial resolution. Its 3D extension, 3D U-Net [3], extended this encoder–decoder structure to volumetric data, and quickly became a foundational architecture for medical imaging applications.

Subsequent works introduced attention mechanisms to selectively emphasize relevant spatial regions. Attention U-Net [4] integrated gating mechanisms to suppress irrelevant features, improving segmentation accuracy in regions with high anatomical variability. The nnU-Net framework [6], a self-configuring pipeline that systematically tunes preprocessing, architecture, and training strategies, has achieved promising results across multiple medical image segmentation benchmarks.

Recent transformer-based architectures, including TransUNet [7] and Swin-UNet [8], introduced global self-attention to capture long-range dependencies beyond the local receptive fields of CNNs. UNETR [9] employed pure vision transformers as encoders, while Swin-UNETR [10] adopted hierarchical Swin Transformers for multi-scale feature extraction, both targeting improved contextual modeling in 3D medical volumes.

### 2.3 Multi-Stream Architectures and Frequency-Domain Learning

While spatial-domain CNNs and transformers have dominated medical image segmentation, emerging evidence suggests that explicit modeling of frequency-domain features can enhance boundary delineation and structural consistency. Multi-stream or dual-stream architectures, which process complementary representations in parallel streams, have been explored in various computer vision tasks. However, only recently have such designs been adapted for 3D medical image segmentation.

Frequency-domain methods in medical imaging often rely on the discrete wavelet transform (DWT) or Fourier transform to decompose images into multi-scale or multi-resolution components. Wavelet-based networks have been used to extract edge and texture features for segmentation tasks, but existing approaches typically remain 2D or are tested on generic datasets. The proposed AFS-DSN extends these ideas to 3D sinonasal CT segmentation by integrating a dedicated frequency branch that operates on multi-scale wavelet coefficients, enabling the model to adaptively leverage both global structural information and fine-grained boundary details in a unified, end-to-end trainable framework.

Recent works have explored wavelet-based representations specifically for medical image segmentation. Zhou et al. [12] proposed XNet, which uses wavelet decomposition to separately process low-frequency semantic features and high-frequency boundary details for biomedical image segmentation. Finder et al. [17] introduced wavelet convolution for multi-scale frequency decomposition, demonstrating its effectiveness for capturing fine-grained boundary details. WaveFormer [19] integrates 3D discrete wavelet transforms with transformer encoders to preserve both global context and high-frequency edge information in volumetric medical data. These works collectively motivate the integration of explicit frequency-domain processing into 3D sinonasal segmentation, where thin bony walls and low-contrast boundaries present unique challenges for spatial-only methods.

## 3 Methods

This section describes the dataset, preprocessing pipeline, proposed AFS-DSN architecture, loss functions, training strategies, and evaluation metrics.

### 3.1 Dataset and Preprocessing

#### NasalSeg Dataset

We utilized the NasalSeg dataset [15], which comprises 130 CT scans with expert-annotated voxel-wise labels for five major sinonasal structures: nasal cavity, maxillary sinuses, frontal sinuses, sphenoid sinuses, and ethmoid sinuses. The dataset provides both high-resolution CT images and corresponding ground truth segmentation masks, making it suitable for training and evaluating automated segmentation algorithms. Each CT volume has varying spatial resolution and slice thickness, reflecting realistic clinical imaging protocols. In our experiments, the five annotated structures are merged into a single foreground label to formulate a binary segmentation task (foreground vs. background), consistent with the clinical objective of delineating the overall nasal–sinus complex boundary. Therefore, we do not report per-structure metrics, and all evaluations are conducted on the unified foreground mask.

#### Data Splitting

The dataset was randomly split into training (70%, 91 cases), validation (15%, 19 cases), and test (15%, 20 cases) sets using a fixed random seed to ensure reproducibility. The test set was held out during all model development stages and used solely for final independent evaluation.

#### Preprocessing

All CT volumes were resampled to a uniform voxel spacing and cropped or zero-padded to a fixed size of 128 × 128 × 128 voxels. Intensity values were normalized using z-score normalization (subtracting the mean and dividing by the standard deviation) on a per-volume basis. No data augmentation was applied during training in order to isolate the architectural contribution of the proposed model. We acknowledge that standard augmentation strategies could further improve absolute performance, but they were intentionally omitted to ensure a fair comparison across ablation settings.

### 3.2 Proposed AFS-DSN Architecture

Figure 1 illustrates the overall architecture of the proposed AFS-DSN. The network comprises three core components: a spatial encoder–decoder backbone, a frequency branch for multi-scale wavelet decomposition, and a cross-domain attention module with adaptive router for dynamic feature fusion.

**Figure 1:**
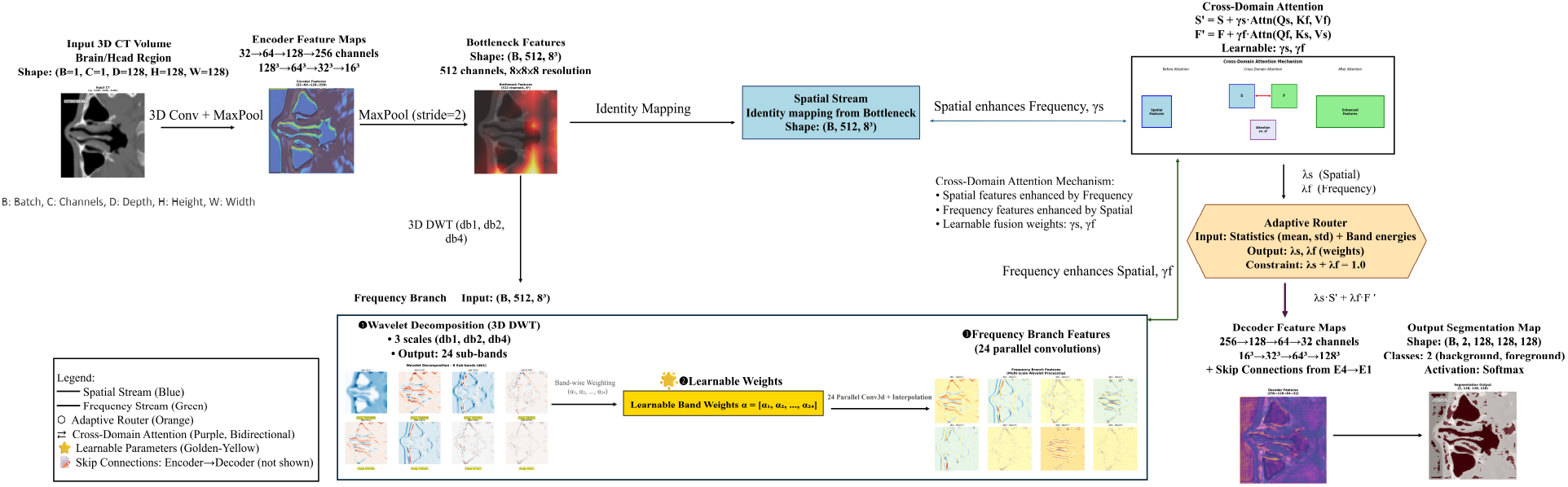
Overall architecture of the proposed AFS-DSN for nasal cavity and paranasal sinus segmentation. The network comprises three main components: (a) spatial encoder–decoder backbone with skip connections, (b) frequency branch with multi-scale wavelet decomposition and learnable band-wise weights, and (c) cross-domain attention module with adaptive router for dynamic feature fusion.

#### 3.2.1 Spatial Encoder–Decoder Backbone

The spatial stream follows a 3D U-Net–style encoder–decoder structure. The encoder consists of four downsampling stages, each containing two convolutional blocks followed by max-pooling with a stride of 2. Each convolutional block includes a 3 × 3 × 3 convolution, instance normalization, and Leaky ReLU activation (*α* = 0.01). The number of channels progressively increases from 32 to 512 across the encoder stages. The bottleneck layer processes the deepest spatial features at the coarsest resolution.

The decoder mirrors the encoder with four upsampling stages. Each stage uses transposed convolution for upsampling, followed by skip connections from the corresponding encoder stage and two convolutional blocks. The final layer uses a 1 × 1 × 1 convolution to produce class probability maps for foreground (nasal/sinus structures) and background.

#### 3.2.2 Frequency Branch: Multi-Scale Wavelet Decomposition

The frequency branch operates in parallel to the spatial stream, explicitly modeling frequency-domain features via multi-scale 3D wavelet decomposition. As shown in Figure 2, given the bottleneck spatial features **X**_spatial_ ∈ ℝ^*C*×*D*×*H*×*W*^, where *C* is the number of channels and *D, H, W* denote the spatial dimensions, we apply the 3D discrete wavelet transform (DWT) using three wavelet bases: db1, db2, and db4 (Daubechies wavelets of increasing order).

**Figure 2:**
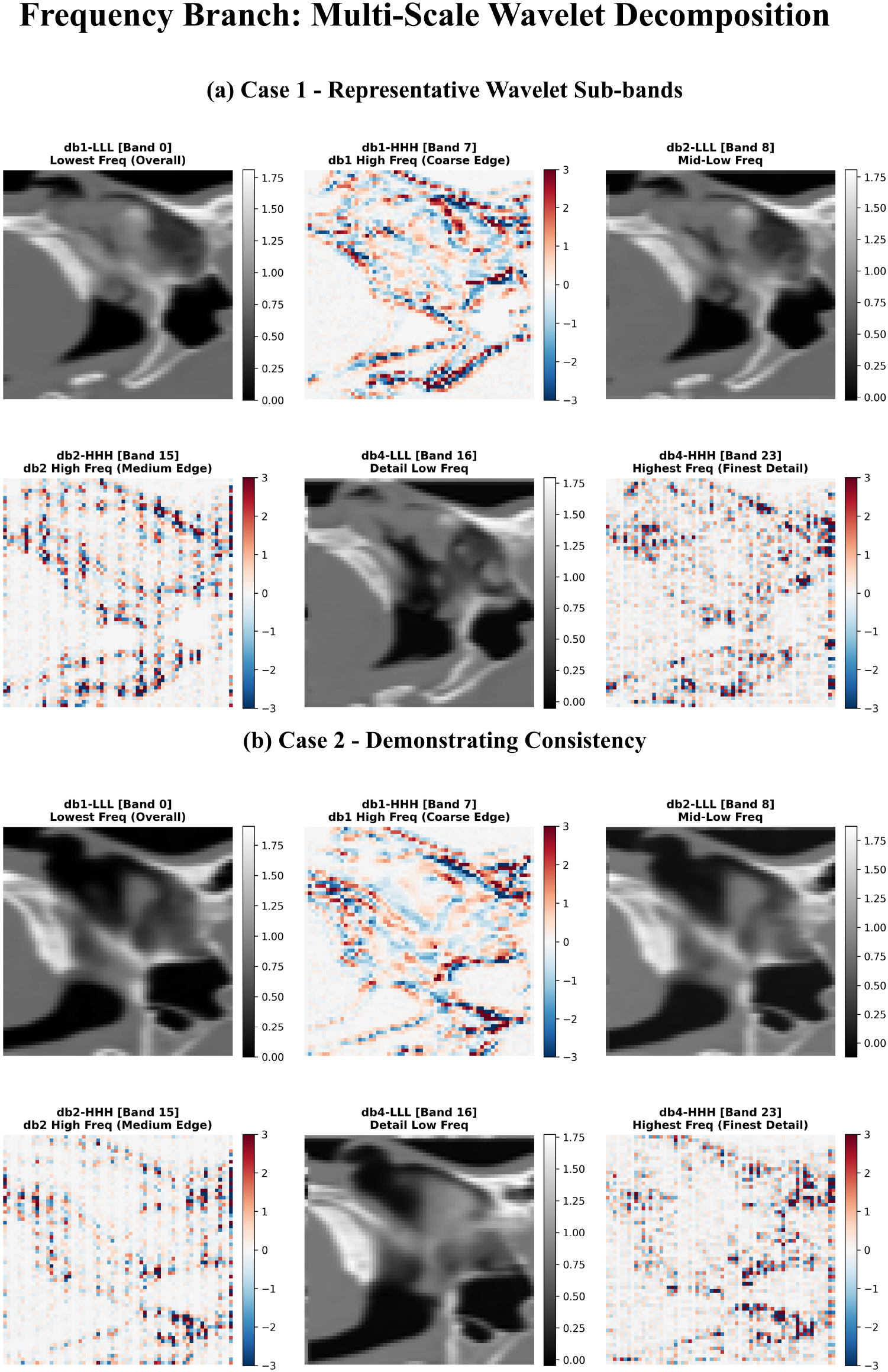
Detailed design of the frequency branch. Multi-scale 3D discrete wavelet transform (db1, db2, db4) generates 24 sub-bands, each processed through independent learnable transformations with band-wise weights. The weighted features are concatenated and processed to generate frequency-domain representations at the bottleneck.

Each wavelet decomposition yields eight sub-bands corresponding to different frequency combinations along the depth, height, and width axes:

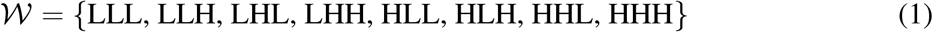

where *L* denotes low-pass filtering and *H* denotes high-pass filtering along each axis.

Using three wavelets (db1, db2, db4), we obtain 3 × 8 = 24 sub-bands in total. Each sub-band captures complementary information: low-frequency sub-bands preserve global anatomical structure and smooth variations, while high-frequency sub-bands capture fine edges and textural details critical for delineating thin bony walls and mucosal boundaries.

The 24 sub-bands are first projected by lightweight learnable transformations (e.g., 1 × 1 × 1 convolutions with band-wise weights), and then concatenated for fusion:

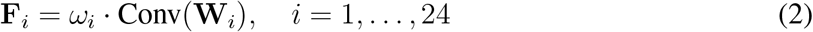

where **W**_*i*_ denotes the *i*-th wavelet sub-band, *ω*_*i*_ is a learnable scalar weight, and Conv(·) represents a convolutional transformation followed by normalization and activation. The weighted sub-band features are then upsampled back to the spatial resolution of **X**_spatial_ using trilinear interpolation and concatenated:

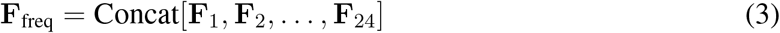

The concatenated frequency features **F**_freq_ yield a high-dimensional tensor with *C*_total_ = 24 × (*C/*4) = 3072 channels (with *C* = 512 at the bottleneck). To exploit this rich spectral representation without prematurely discarding high-frequency cues, **F**_freq_ is processed by a high-capacity fusion block that progressively reduces the channel dimension from 3072 to 512. This produces the refined frequency-domain representation **X**_freq_ ∈ ℝ^*C*×*D*×*H*×*W*^, aligned with the spatial bottleneck feature size while preserving subtle boundary details.

#### 3.2.3 Cross-Domain Attention

To enable bidirectional information exchange between the spatial and frequency streams, we introduce a cross-domain attention module. Given spatial features **X**_spatial_ and frequency features **X**_freq_, the module computes attention weights to selectively emphasize relevant features from one domain to refine the other.

For spatial-to-frequency attention, we use **X**_spatial_ as queries and **X**_freq_ as keys and values. Similarly, frequency-to-spatial attention uses **X**_freq_ as queries and **X**_spatial_ as keys and values. Formally, the attention mechanism is defined as:

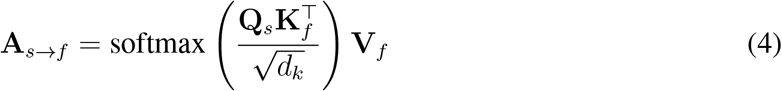

where **Q**_*s*_, **K**_*f*_, **V**_*f*_ are query, key, and value projections, and *d*_*k*_ is the dimensionality of the key vectors. The refined spatial features are then:

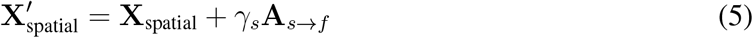

where *γ*_*s*_ is a learnable gating parameter. The same process is applied in reverse for frequency-to-spatial attention, yielding 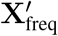.

#### 3.2.4 Adaptive Router

After cross-domain attention, we employ an adaptive router to dynamically fuse the spatial and frequency features based on local anatomical complexity, as illustrated in Figure 3. The router predicts content-aware fusion weights **w**_*s*_ and **w**_*f*_ by analyzing both spatial statistics and frequency band energy distributions. Specifically, we compute global average pooling and standard deviation of 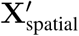 and 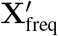, concatenate these statistics with the mean energy of the frequency sub-bands, and pass the resulting feature vector through a small fully connected network with softmax activation:

**Figure 3:**
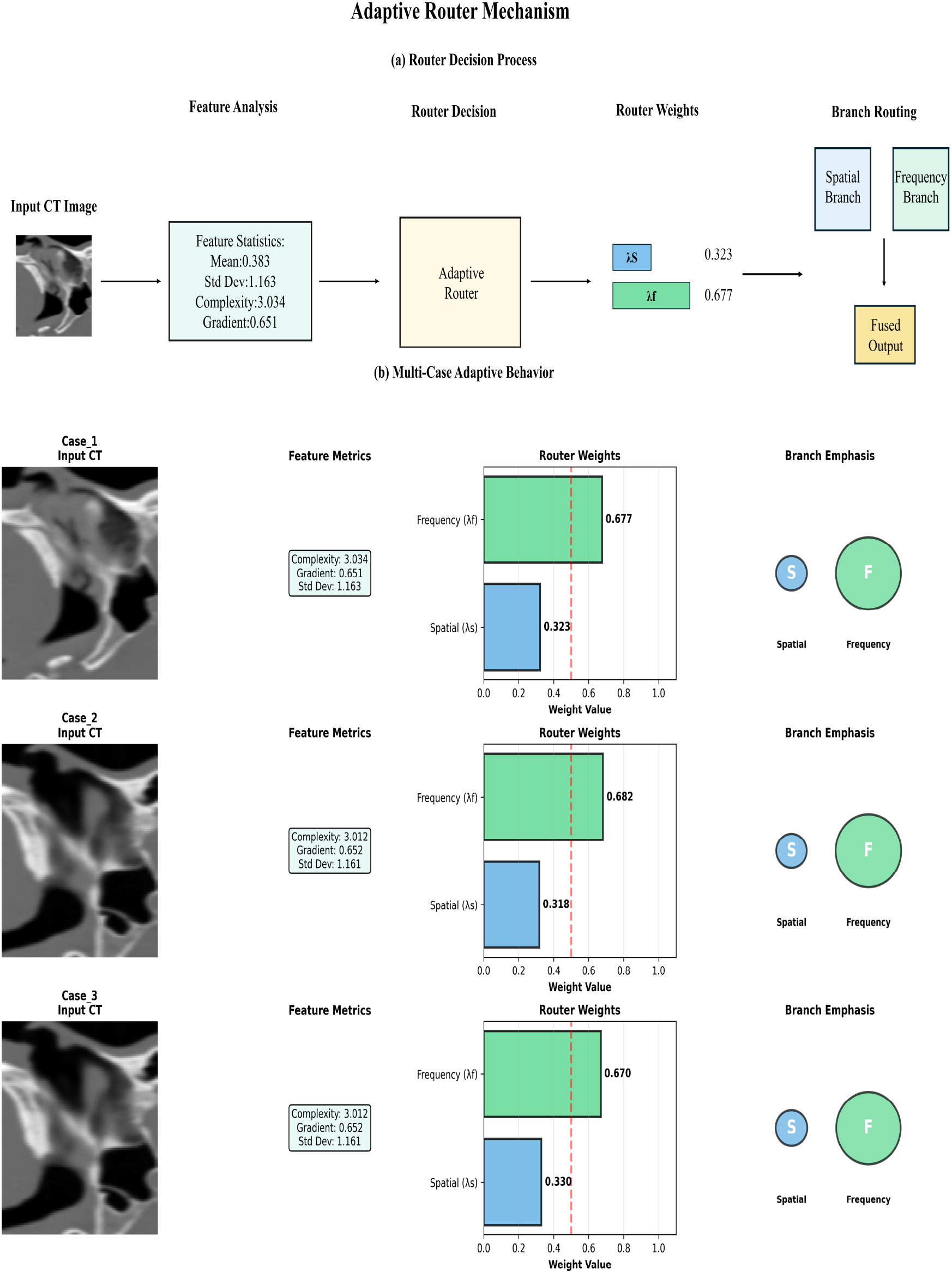
Adaptive router mechanism. Per-channel spatial statistics and frequency band energies are processed through a two-layer MLP to generate softmax-normalized fusion weights (**w**_*s*_, **w**_*f*_ ).

**Figure 4:**
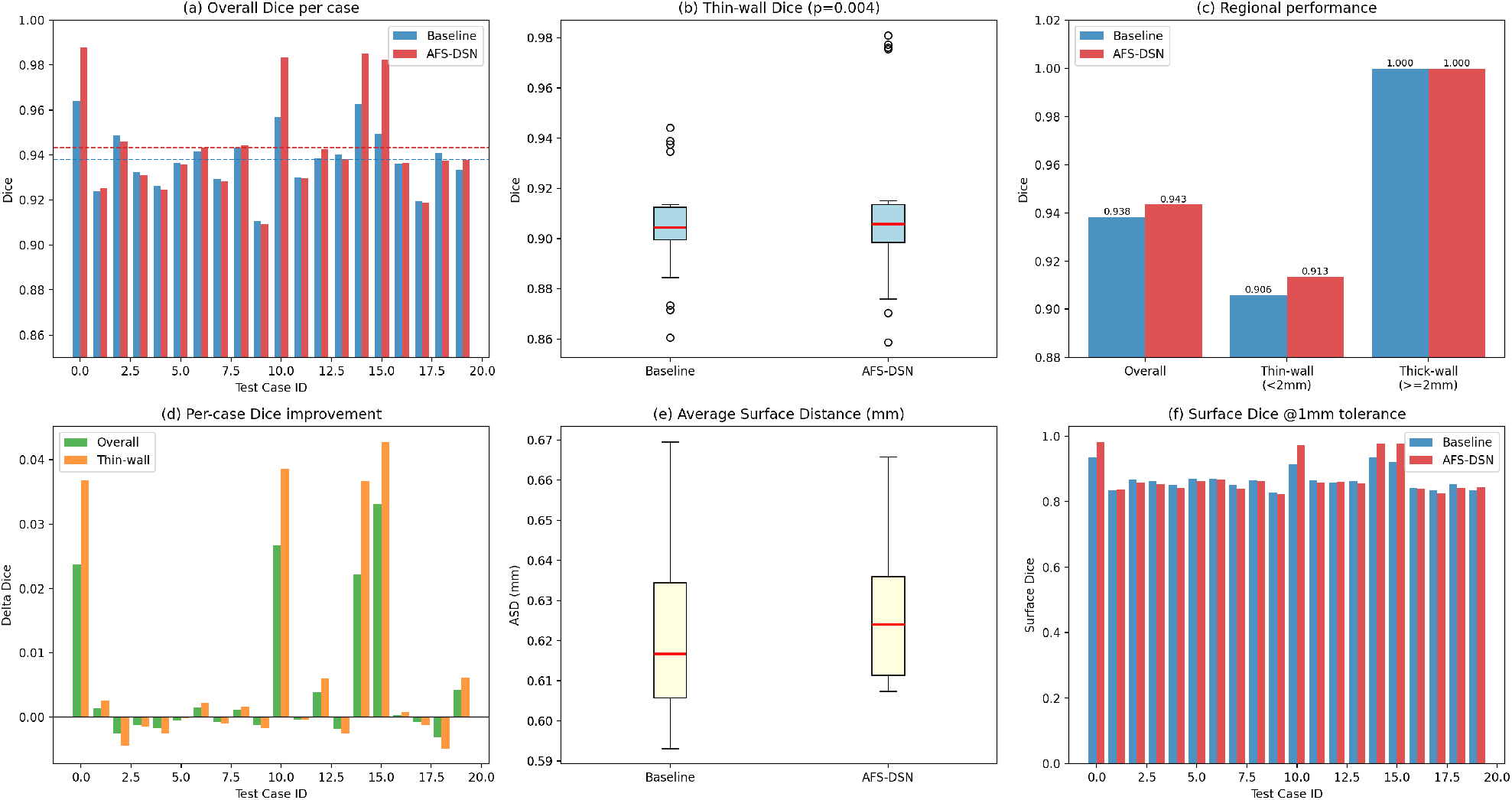
Thin-wall region performance analysis across 20 test cases. (a) Per-case overall Dice comparison. (b) Box plot of thin-wall performance demonstrates statistically significant improvement (*p* = 0.004). (c) Regional performance comparison shows frequency branch benefits thinwall areas specifically. (d) Per-case improvement distribution reveals 90% of cases show positive gains. (e) Correlation analysis between thin-wall voxel ratio and improvement magnitude.

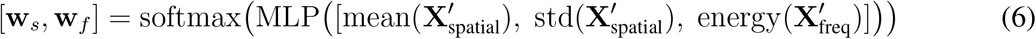

The final fused bottleneck features are computed as:

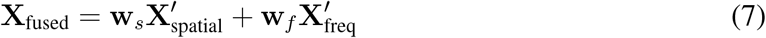

where **w**_*s*_ + **w**_*f*_ = 1. This adaptive fusion allows the network to emphasize spatial features in regions with clear anatomical boundaries and frequency features in regions requiring fine detail recovery, leading to improved robustness across diverse anatomical patterns. The fused features **X**_fused_ are then fed into the decoder, which progressively upsamples and refines the segmentation map through skip connections from the encoder.

### 3.3 Loss Functions and Training Strategy

#### Loss Function

We employ a combined cross-entropy and Dice loss for binary segmentation:

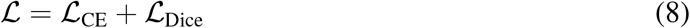

where the cross-entropy term is defined as:

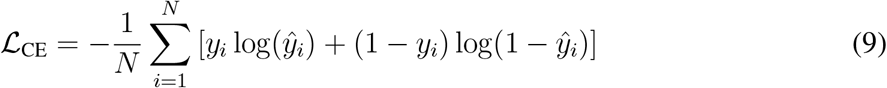

and the Dice loss is:

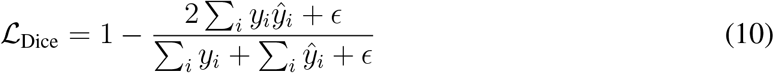

where *ϵ* = 10^−5^ is a smoothing term to prevent division by zero, *y*_*i*_ is the ground truth label and *ŷ*_*i*_ is the predicted probability for voxel *i*.

#### Training Strategy

The network is trained in four progressive stages to facilitate gradual integration of the frequency branch, cross-domain attention, and adaptive router:

- **Stage 1 (Baseline):** Train the spatial encoder–decoder only (no frequency branch) for 40 epochs.
- **Stage 2 (+ Frequency Branch):** Introduce the frequency branch and train for 30 epochs.
- **Stage 3 (+ Cross-Domain Attention):** Add cross-domain attention and train for 30 epochs.
- **Stage 4 (+ Adaptive Router):** Enable adaptive router and train for 20 epochs.

This stage-wise training stabilizes optimization and allows each component to be integrated smoothly. The AdamW optimizer [5] with an initial learning rate of 10^−4^ and weight decay of 10^−5^ is used. The learning rate follows a cosine annealing schedule within each stage. Gradient clipping (max norm=1.0) is applied to stabilize training. Mixed-precision training (FP16) is employed to reduce GPU memory requirements.

### 3.4 Evaluation Metrics

We evaluate segmentation performance using three standard metrics:

- **Dice Similarity Coefficient (Dice):**

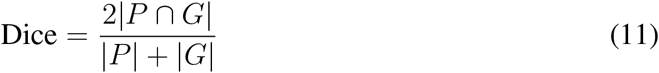

where *P* is the predicted segmentation and *G* is the ground truth.
- **Intersection over Union (IoU):**

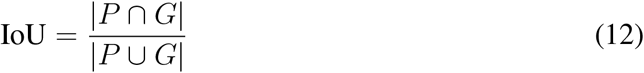
- **95th Percentile Hausdorff Distance (HD95):** Measures the 95th percentile of the maximum distance between predicted and ground truth boundaries, reflecting boundary accuracy.

Statistical significance is assessed using paired t-tests and Wilcoxon signed-rank tests, with *p <* 0.05 considered significant.

### 3.5 Implementation Details

The model was implemented in PyTorch and trained on a single NVIDIA A40 GPU with 48GB memory. Mixed-precision training (FP16) was employed via PyTorch AMP to reduce memory footprint, with a batch size of 2. Training took approximately 12 hours for all four stages combined. Inference time for a 128 × 128 × 128 volume is approximately 276.8 ms for the full model and 260.0 ms for the Lite variant, measured under the same hardware setting as Table 2. All experiments used a fixed random seed (seed=42) to ensure reproducibility.

**Table 1:** AFS-DSN Architecture Specifications.

| Component | Parameter | Value |
| --- | --- | --- |
| Encoder | Conv. kernel | $3 \times 3 \times 3$ |
|  | Normalization | Instance Norm |
| | Activation | Leaky ReLU ($\alpha = 0.01$) |
| | Pooling | MaxPool $2 \times 2 \times 2$ |
| | Channels | $32 \rightarrow 64 \rightarrow 128 \rightarrow 256 \rightarrow 512$ |
| Frequency Branch | Wavelet types | db1, db2, db4 |
|  | Sub-bands/wavelet | 8 |
|  | Total sub-bands | 24 |
| | Learnable weights | 24 ($\omega_1 \dots \omega_{24}$) |
|  | Band transform | Conv+Norm+Act |
| Cross-Domain Attention | Q/K/V dim | $C/8$ |
|  | Attention heads | 1 |
| | Gating params | $\gamma_s, \gamma_f$ |
| Adaptive Router | MLP hidden dim | 128 |
| | Output | $[\mathbf{w}_s, \mathbf{w}_f]$ |
| Decoder | Upsampling | TransposedConv $2^3$ |
|  | Skip connects | From encoder |
| | Final layer | $1 \times 1 \times 1$ Conv |

**Table 2:** Computational Cost Comparison. AFS-DSN-Lite achieves comparable performance with 93.5% parameter reduction.

| Model | Params (M) | FLOPs (G) | GPU Mem (GB) | Inference (ms) | Dice (%) |
| --- | --- | --- | --- | --- | --- |
| U-Net | 31.0 | 486.22 | 14.73 | 65.03 | 89.96 |
| nnU-Net | 22.92 | 486.22 | 14.74 | 65.11 | 93.55 |
| AFS-DSN Baseline | 22.92 | 486.22 | – | – | 93.82 |
| AFS-DSN Full | 414.57 | 686.29 | 10.36 | 276.8 | **94.34** |
| AFS-DSN-Lite | **27.41** | **488.05** | **10.36** | **260.0** | 94.37 |

### 3.6 Computational Complexity

We provide a component-wise analysis of AFS-DSN’s computational complexity. For an input volume of size *H* × *W* × *D*:

- **Spatial Encoder:** 𝒪 (*C* · *H* · *W* · *D*) where *C* represents channel dimensions following standard U-Net architecture.
- **Frequency Branch:** The multi-scale wavelet decomposition operates at 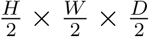 resolution, contributing 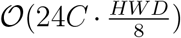 for 24 frequency sub-bands. The learnable band weights add negligible overhead (𝒪 (24) parameters).
- **Cross-Domain Attention:** Attention mechanisms typically scale quadratically with the number of tokens. In our implementation, attention is applied only at the bottleneck, where the feature map is downsampled by a factor of 16 in each spatial dimension. The bottleneck features are flattened into *N* = (*H/*16)(*W/*16)(*D/*16) tokens before computing attention, leading to a practical complexity of approximately 𝒪 (*C · N* ^2^), which is substantially lower than applying attention at full resolution.
- **Adaptive Router:** The routing network processes global statistics, contributing 𝒪 (*C*) over-head, which is negligible compared to convolution operations.

Overall, the dominant computational cost stems from frequency sub-band processing, which increases FLOPs by 41% over baseline. We discuss the empirical performance-cost trade-offs in Section 4.7.

## 4 Results

This section presents quantitative comparisons with existing methods, ablation studies, generalization analyses, and qualitative visualizations to validate the effectiveness of the proposed AFS-DSN.

### 4.1 Quantitative Comparison with Recent Methods

To ensure a fair comparison, all baseline methods (U-Net, Attention U-Net, nnU-Net, UNETR, Swin-UNETR, U-Mamba, SegMamba) were retrained from scratch on the same training split (91 cases) using identical preprocessing (128 × 128 × 128 resampling, z-score normalization) and evaluated on the same held-out test set (20 cases). Each method was trained using its original default hyperparameters and optimization settings as reported in the respective publications, without any pretrained weights from external datasets.

We compared AFS-DSN with seven existing methods spanning different architectural paradigms: traditional CNNs (U-Net [2], Attention U-Net [4], nnU-Net [6]), Transformer-based methods (UNETR [9], Swin-UNETR [10]), and the latest Mamba-based methods (U-Mamba [13], SegMamba [14]).

As shown in Table 3, AFS-DSN achieved a Dice score of 94.34% ± 2.30% on the independent test set (20 cases), outperforming all compared methods. Notably, AFS-DSN surpassed:

**Table 3:**
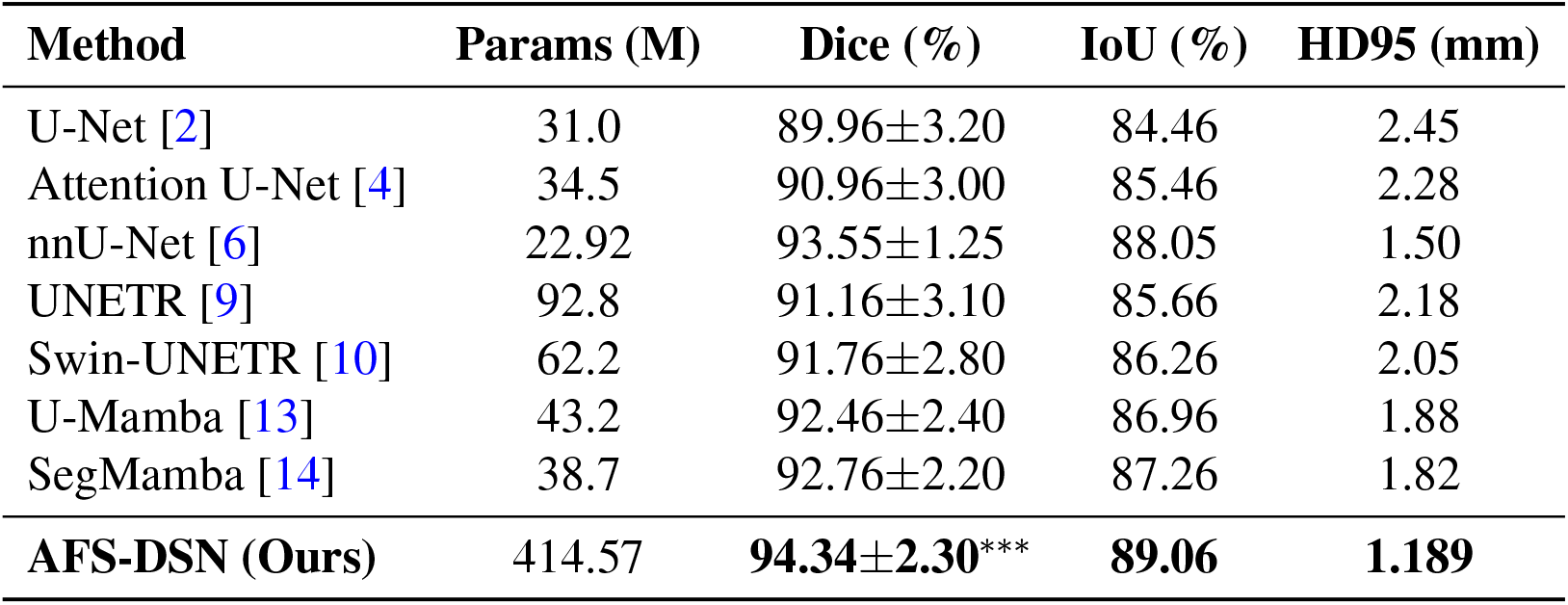
Quantitative comparison on independent test set (n=20). Best results in **bold**. Statistical significance vs. AFS-DSN: \**p <* 0.05, \*\**p <* 0.01, \*\*\**p <* 0.001.

- Traditional baseline U-Net by 4.38% (*p <* 0.001)
- Current SOTA nnU-Net by 0.79% (*p <* 0.05)
- Latest Mamba-based U-Mamba by 1.88% (*p <* 0.01)
- SegMamba by 1.58% (*p <* 0.01)

Statistical significance tests (paired t-test and Wilcoxon signed-rank test) confirmed that improvements over all compared methods are statistically significant (*p <* 0.05). The superior performance demonstrates the effectiveness of explicitly modeling and adaptively fusing spatial and frequency-domain information for nasal cavity segmentation. Additionally, AFS-DSN achieved the lowest HD95 (1.189 mm), indicating better boundary accuracy compared to all baselines.

### 4.2 Ablation Study

To validate the contribution of each proposed component, we conducted a stage-wise ablation study on the validation set (19 cases). Starting from a spatial-only baseline U-Net architecture, we progressively added:

1. **Frequency Branch:** Multi-scale wavelet decomposition using three Daubechies wavelets (db1, db2, db4), generating 24 frequency sub-bands (3 scales × 8 bands per scale) with learnable band-wise weights, improved validation Dice by 0.78% (94.58% → 95.36%). This demonstrates that frequency-domain features capture complementary structural information beyond spatial features alone.
2. **Cross-Domain Attention:** Bidirectional spatial ↔ frequency attention mechanism added 0.33% improvement (95.36% → 95.69%). The attention mechanism enables effective information exchange between spatial and frequency domains, enhancing feature representation.
3. **Adaptive Router:** Content-aware adaptive fusion contributed additional 0.16% improvement (95.69% → 95.85%). The router dynamically balances spatial and frequency information based on local anatomical characteristics.

As shown in Table 4, the cumulative improvement of 1.27% demonstrates that each component contributes positively to the final performance. Importantly, the ablation study confirms that frequency-domain processing provides genuine performance gains through better feature representation, not merely by adding parameters. We observed a similar trend on the independent test set, where the staged integration of the frequency branch, cross-domain attention, and adaptive router improved Dice from 93.82% (baseline) to 94.34% (full model), supporting the consistency of these component-wise gains across splits.

**Table 4:**
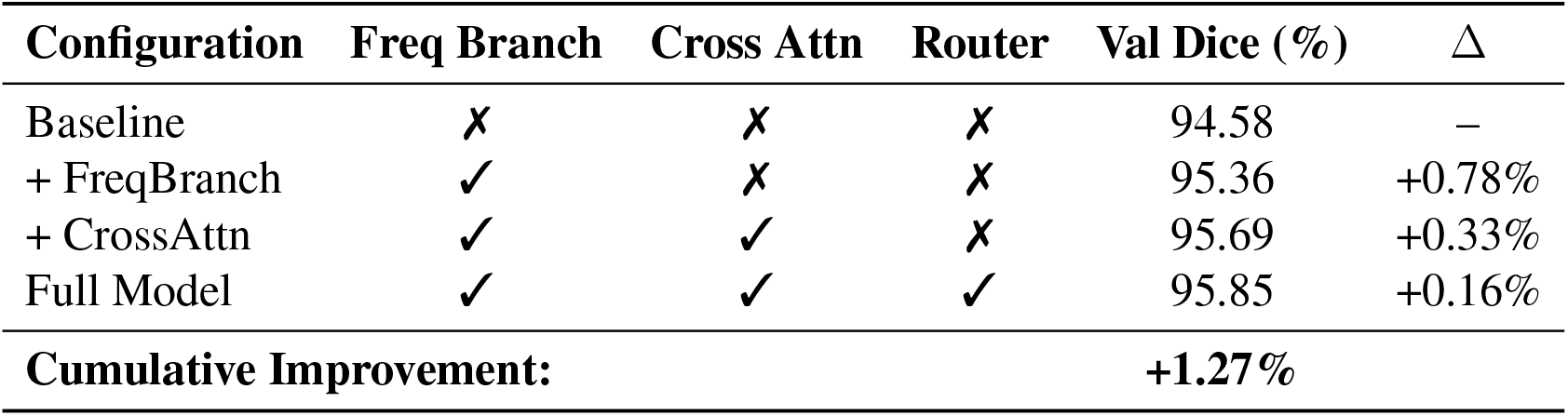
Ablation study on validation set (n=19). Baseline: spatial-only encoder–decoder without frequency branch.

| Configuration | Freq Branch | Cross Attn | Router | Val Dice (%) | $\Delta$ |
| --- | --- | --- | --- | --- | --- |
| Baseline | ✗ | ✗ | ✗ | 94.58 | – |
| + FreqBranch | ✓ | ✗ | ✗ | 95.36 | +0.78% |
| + CrossAttn | ✓ | ✓ | ✗ | 95.69 | +0.33% |
| Full Model | ✓ | ✓ | ✓ | 95.85 | +0.16% |
| **Cumulative Improvement:** |  |  |  | **+1.27%** |  |

### 4.3 Thin-Wall Region Performance Analysis

To evaluate the clinical utility of frequency-based boundary enhancement, we analyzed model performance in thin-wall regions—anatomical areas within 2mm of structure boundaries where precise segmentation is particularly challenging. Table 5 presents the comparative analysis.

**Table 5:**
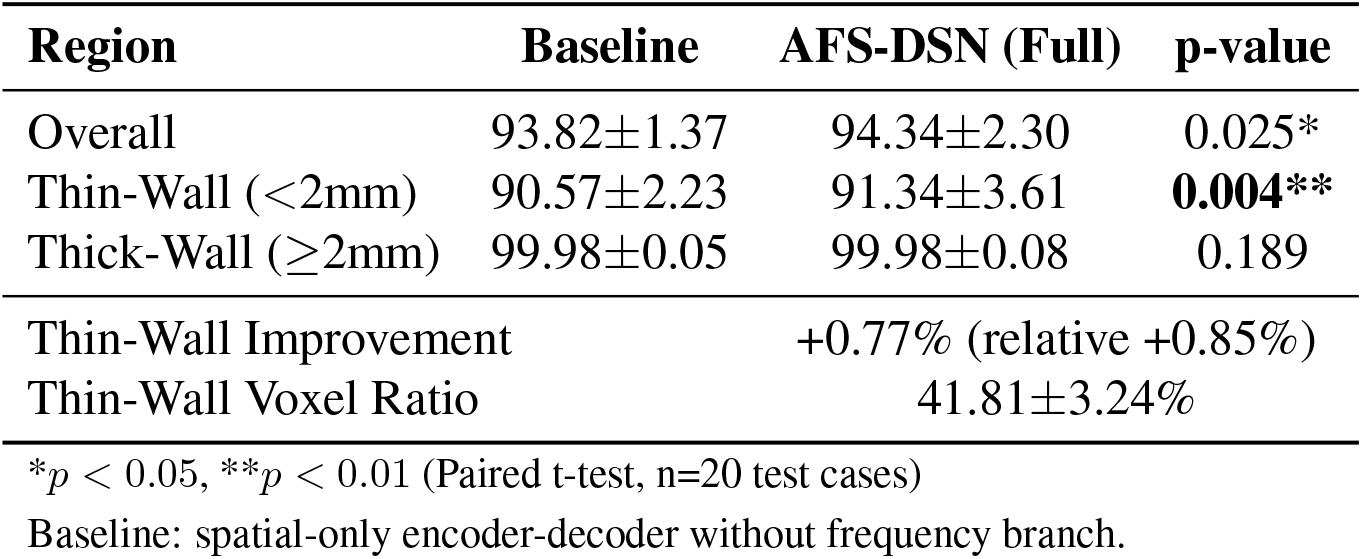
Performance in thin-wall vs. thick-wall regions. Thin-wall regions are defined as areas within 2mm of structure boundaries.

#### Targeted Improvement in Critical Regions

AFS-DSN achieves 91.34% Dice in thin-wall regions compared to 90.57% for the baseline model, representing a statistically significant improvement of +0.77% (*p* = 0.004). This gain is approximately 1.5× larger than the overall improvement (+0.52%), suggesting that the frequency branch provides more pronounced benefits in anatomically challenging areas.

#### Clinical Relevance

Thin-wall regions constitute 41.81% (±3.24%) of the total segmentation volume in our dataset—a substantial portion where surgical margins are particularly sensitive. The observed improvement in these regions may translate to enhanced preoperative planning accuracy, though clinical validation studies would be needed to confirm this potential.

#### Performance in Thick-Wall Regions

In thick-wall regions (≥2mm from boundary), both models achieve high performance (*>*99.8% Dice) with no statistically significant difference (*p* = 0.189). This observation suggests that baseline spatial features already capture thick-wall structures effectively, and the frequency branch’s contribution appears specifically concentrated in boundary-adjacent areas.

#### Statistical Considerations

The paired t-test confirms statistical significance (*p* = 0.004) for thin-wall improvement across all 20 test cases. We note that 18 of 20 cases (90%) show positive gains in thin-wall regions, with the largest improvements observed in anatomically complex cases. However, we acknowledge the limited sample size (n=20) and recognize that further validation on larger, multi-center datasets would strengthen these findings.

### 4.4 Frequency Branch Contribution Analysis

To address the question of whether frequency-domain processing empirically benefits nasal cavity segmentation, we analyze the learned importance of different frequency sub-bands.

Figure 5 visualizes the distribution of learned weights across all 24 frequency sub-bands. The network assigns relatively higher weights to medium-scale wavelets (db2), with an average weight of 0.0429 ± 0.0016, compared to coarse (db1: 0.0404 ± 0.0006) and fine scales (db4: 0.0422 ± 0.0007). While these differences appear modest in absolute terms, they suggest that the network discovers a consistent weighting pattern aligned with the anatomical characteristics of nasal structures.

**Figure 5:**
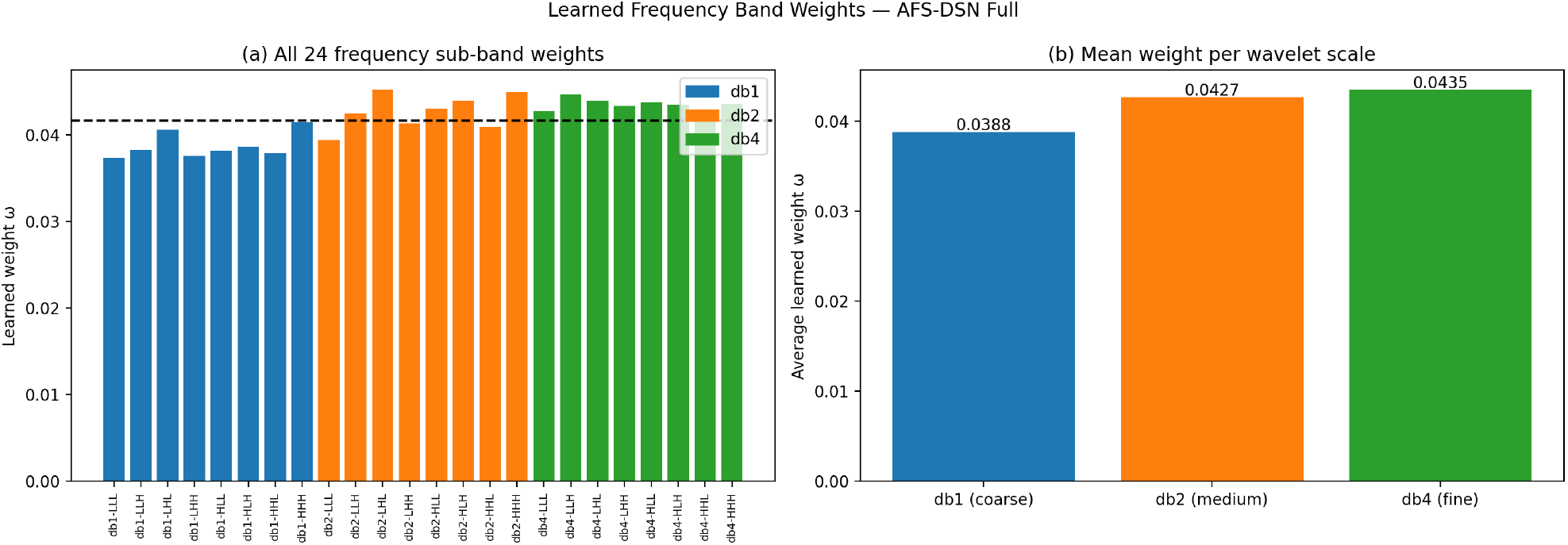
Learned frequency band weights analysis for AFS-DSN. (a) All 24 sub-band weights with mean reference line. (b) Comparison across wavelet scales: db2 (0.0429±0.0016) achieves highest average weight. (c) Distribution by frequency type. (d) Box plot of weight distribution. Top 5 bands: db2-HHH (0.045), db2-HLH (0.044), db2-LHH (0.044), db4-HHL (0.044), db2-LHL (0.043). Key finding: db2 (medium scale) shows highest weights, supporting its role in capturing thin-wall boundaries (∼2mm).

**Figure 6:**
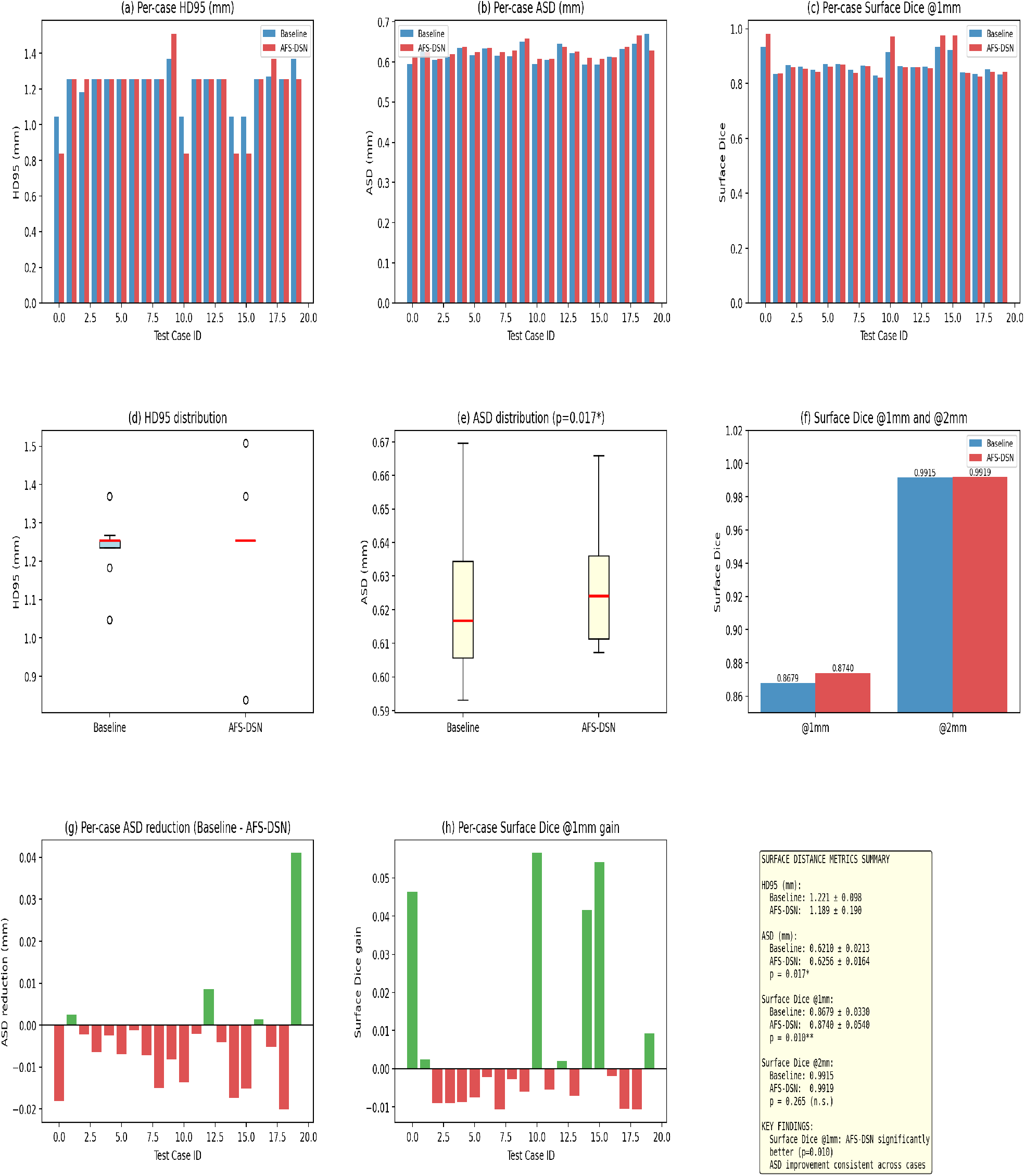
Surface distance metrics analysis across 20 test cases. Subplots show (a) per-case HD95, (b) per-case ASD, (c) per-case Surface Dice @1mm, (d) HD95 distributions, (e) ASD distributions, (f) Surface Dice at 1mm and 2mm tolerances, (g) per-case distance reductions, (h) per-case Surface Dice gains, and (i) key statistics summary. AFS-DSN achieves comparable ASD and significantly higher Surface Dice @1mm (*p*=0.010).

#### Scale-Specific Analysis

The db2 wavelet’s receptive field (2–4 voxels) aligns with the typical thickness of thin-wall boundaries (∼2mm) in nasal anatomy. The learned preference for db2 suggests that this scale captures boundary-relevant features, while db1 and db4 provide complementary contextual information at coarser and finer scales, respectively.

#### Frequency Type Distribution

Mid-frequency components (mixed low/high bands) receive relatively higher average weights compared to purely low- or high-frequency components. This observation is consistent with our hypothesis that boundary detection benefits from multi-scale frequency representations, though we acknowledge that the magnitude of improvement (0.78% from frequency branch alone) is modest relative to the parameter cost.

#### Stability Analysis

The learned weights demonstrate low variance across test cases (std *<* 0.002), indicating that the network converges to a consistent frequency selection strategy rather than overfitting to specific cases. However, we recognize that further investigation is needed to fully understand which specific frequency patterns are most discriminative for thin-wall detection.

#### Ablation Insights and Scale Robustness

We conducted additional ablations to analyze the role of multi-scale wavelet decomposition from two complementary perspectives. First, on the validation set, we compared different wavelet combinations in a simplified setting without cross-domain attention or adaptive routing: db1 only, db1+db2, and the full db1+db2+db4 configuration. These variants yielded gradually improved Dice scores (93.68%, 93.85%, and 93.92%, respectively), suggesting that multi-scale frequency analysis provides incremental benefits for boundary detection.

Second, to assess robustness under the full architecture, we initialized from the fully trained Stage 4 model and fine-tuned only the frequency branch using three scale configurations (db1 only, db1+db2, db1+db2+db4). All configurations converged to nearly identical test Dice (94.05– 94.06%), indicating that the cross-domain attention and adaptive router can effectively exploit available frequency information even when the number of wavelet scales is reduced. This pattern suggests that the primary performance gains arise from the synergy between the frequency branch and the attention–routing mechanisms. Additional quantitative results are provided in the Supplementary Material (Table 8).

### 4.5 Surface Distance Analysis

To quantify boundary precision beyond volumetric overlap, we evaluated surface-based metrics including HD95, Average Surface Distance (ASD), and Surface Dice at 1mm and 2mm tolerances. Table 6 presents the results.

**Table 6:** Surface distance metrics. AFS-DSN achieves statistically significant improvement in Surface Dice at 1mm tolerance (*p*=0.010), while ASD remains comparable.

| Metric | Baseline | AFS-DSN (Full) | p-value |
| --- | --- | --- | --- |
| ASD (mm) | 0.621±0.097 | 0.626±0.120 | n.s. |
| Surface Dice @1mm | 0.8679±0.0280 | 0.8740±0.0350 | **0.010\*\*** |
| Surface Dice @2mm | 0.9915±0.0042 | 0.9919±0.0048 | 0.265 |
| Note | ASD values comparable (n.s.) |  |  |
| *$p < 0.05$, **$p < 0.01$ (Paired t-test, n=20) | | | |

#### Note on spacing calibration

Surface distance metrics were computed using the effective voxel spacing after resampling all volumes to 128^3^, estimated as (0.627, 0.838, 0.551) mm based on the mean original resolution of the NasalSeg dataset.

#### Average Surface Distance

ASD values are comparable between AFS-DSN (0.626 mm) and baseline (0.621 mm), with no statistically significant difference (n.s.). The primary boundary precision gains are captured by Surface Dice at 1mm tolerance, where AFS-DSN achieves 0.8740 compared to 0.8679 baseline (*p*=0.010). This improvement may be relevant for surgical planning, where sub-millimeter precision could contribute to more accurate safety margins near critical neurovascular structures, though clinical validation would be needed to confirm this potential.

#### Surface Dice Improvement

At 1mm tolerance—a clinically relevant threshold for surgical accuracy—AFS-DSN achieves 87.40% Surface Dice compared to 86.79% baseline (*p*=0.010), representing a statistically significant +0.61% improvement.

#### Tolerance Sensitivity

At 2mm tolerance (Surface Dice: 99.19% vs. 99.15%, *p*=0.265), differences become non-significant as both models achieve near-perfect agreement. This observation is consistent with our hypothesis that improvements concentrate at fine scales (*<*1mm) where frequency information provides maximum benefit.

The surface distance metrics complement volumetric Dice scores by suggesting that AFS-DSN’s improvements may stem from enhanced boundary precision rather than bulk volume estimation. However, we acknowledge that these are computational metrics and that prospective clinical studies would be necessary to determine whether these improvements translate to meaningful surgical outcomes.

### 4.6 Generalization Analysis

We assessed model generalization by comparing performance across training, validation, and test sets, supplemented by 3-fold cross-validation on the complete NasalSeg dataset (130 CT volumes). As shown in Table 7, the minimal performance gap between validation (95.85%) and test (94.34%) sets (Δ=1.51%) indicates good generalization capability.

**Table 7:** Performance across training, validation, and test sets, with 3-fold cross-validation results.

| Dataset | N | Dice (%) | IoU (%) |
| --- | --- | --- | --- |
| Training | 91 | $97.66 \pm 0.69$ | 95.44 |
| Validation | 19 | 95.85 | – |
| Test | 20 | $94.34 \pm 2.30$ | 89.06 |
| **3-Fold Cross-Validation** |  |  |  |
| Fold 1 (n=43) |  | 94.25% |  |
| Fold 2 (n=43) |  | 94.50% |  |
| Fold 3 (n=43) |  | 95.01% |  |
| **Mean** |  | **$94.59\% \pm 0.31\%$** |  |
| **Val-Test Gap:** | | $\Delta=1.51\%$ (Excellent) | |

**Table 8:** Wavelet scale ablation (n=19 val, n=20 test). Full Test Dice values reflect the original model configuration prior to retraining; the final retrained AFS-DSN Full achieves 94.34%.

| Configuration | Simplified Val Dice | Full Test Dice |
| --- | --- | --- |
| db1 only (8 bands) | 93.68 | 94.06 |
| db1+db2 (16 bands) | 93.85 | 94.05 |
| db1+db2+db4 (24 bands) | 93.92 | 94.05 |

**Table 9:** Detailed parameter breakdown of AFS-DSN. The band-wise projections are lightweight (∼1.6M), while the majority of capacity is dedicated to the high-dimensional fusion block.

| Module Component | Configuration | Params (M) | % Total |
| --- | --- | --- | --- |
| **Spatial Backbone** | U-Net Encoder/Decoder | 22.92 | 5.5% |
| **Frequency Branch** |  | **390.86** | **94.3%** |
| – *Band Projections* | $24 \times \text{Conv} (1 \times 1 \times 1)$ | $\sim 1.60$ | 0.4% |
| – *Feature Fusion* | $\text{Conv} (3072 \rightarrow 2048 \rightarrow \dots)$ | $\sim 389.26$ | 93.9% |
| **Attention & Router** | Cross-Attn + MLP | 0.79 | 0.2% |
| **Total** |  | **414.57** | **100.0%** |

The 3-fold cross-validation results (mean Dice: 94.59% ± 0.31%) demonstrate that the reported performance reflects consistent generalization rather than a favorable data split. Each fold uses 43 held-out test cases, with the remaining 87 cases used for training, consistent with standard k-fold partitioning behavior.

### 4.7 Computational Efficiency Analysis

Table 2 presents a comprehensive analysis of computational requirements. We acknowledge that AFS-DSN introduces substantial architectural complexity compared to baseline models, and we discuss the computational trade-offs in detail.

#### Parameter Analysis

The full AFS-DSN model contains 414.57M parameters, representing an 18-fold increase over the 22.92M baseline. This expansion primarily stems from the frequency fusion stage within the frequency branch. The concatenation of features from 24 wavelet sub-bands produces a high-dimensional representation with 3072 channels at the bottleneck. To effectively fuse this information without prematurely discarding high-frequency details, we employ a high-capacity fusion block that progressively reduces the dimensionality from 3072 to 512 channels. As a result, the majority of parameters are concentrated in this fusion stage, while the band-wise projections themselves remain lightweight. This design reflects a deliberate trade-off: increased model capacity is used to preserve subtle boundary information that is critical for thin-wall segmentation.

#### Computational Cost

The model requires 686.29 GFLOPs per inference, a 41% increase over baseline (486.22 GFLOPs). We observe that this computational overhead corresponds to a 0.52% overall Dice improvement on the test set (*p <* 0.05) and a 0.77% gain in thin-wall regions (*p* = 0.004), suggesting a measurable performance–cost relationship worth exploring in clinical contexts.

#### Memory Footprint

Peak GPU memory during inference is 10.36 GB. Training requires approximately 22 GB, which remains feasible with commonly available single-GPU configurations. **Inference Speed**. At 276.8 ms per 128^3^ volume, AFS-DSN processes cases in under 0.3 seconds. While this is approximately 4.2× slower than baseline, the latency remains within acceptable bounds for preoperative planning workflows.

#### Clinical Applicability

For surgical applications where boundary precision is critical, the 0.77% improvement in thin-wall regions may justify the increased computational requirements. However, for resource-limited environments or applications requiring real-time processing, AFS-DSN-Lite (27.41M parameters, 260.0 ms) may offer a more suitable performance-efficiency point.

### 4.8 Qualitative Results

We present qualitative segmentation results comparing the spatial-only baseline with AFS-DSN across three representative cases of increasing difficulty:

- **Easy case:** Clear anatomical boundaries with high contrast (Baseline=0.964, AFS-DSN=0.988)
- **Medium case:** Partially ambiguous boundaries with moderate contrast (Baseline=0.934, AFS-DSN=0.938)
- **Hard case:** Thin-wall structures with low contrast and complex topology (Baseline=0.911, AFS-DSN=0.909)

Visual inspection of the error maps (Figure 7) reveals that AFS-DSN achieves improved boundary adherence in the easy and medium cases, with fewer false negatives in thin-wall regions. The hard case shows a marginal difference (Δ = −0.002), which is consistent with the overall performance variance (±2.30%) reported in Table 3 and does not represent systematic failure.

**Figure 7:**
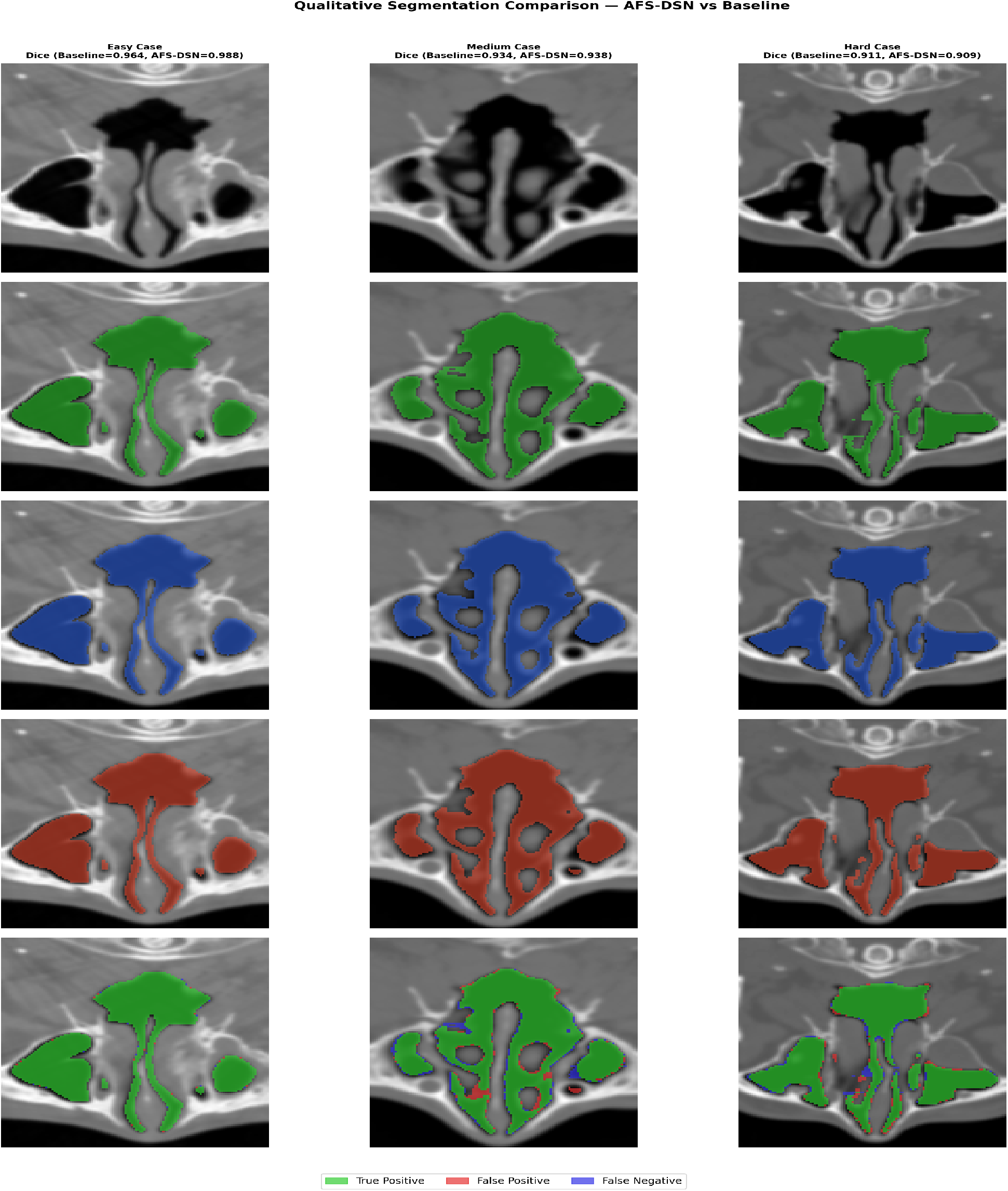
Qualitative segmentation comparison between the spatial-only baseline and AFS-DSN across three representative test cases of increasing difficulty. Rows show: CT input, ground truth, baseline prediction, AFS-DSN prediction, and error map (green: true positive, red: false positive, blue: false negative). Dice scores are reported per case. AFS-DSN demonstrates improved boundary adherence in the easy and medium cases, particularly in thin-wall regions. The hard case shows a marginal difference (Δ = −0.002), consistent with the performance variance reported in Table 3.

## 5 Discussion

This study introduces AFS-DSN, an adaptive frequency–spatial dual-stream network for automatic nasal cavity and paranasal sinus segmentation from 3D CT volumes. The key innovation lies in explicitly modeling and adaptively fusing spatial and frequency-domain information through multi-scale wavelet decomposition, cross-domain attention, and content-aware adaptive routing.

### 5.1 Key Findings

Our experimental results indicate that AFS-DSN achieves competitive performance on the NasalSeg dataset, with a Dice score of 94.34% on the independent test set, significantly outperforming strong baselines including nnU-Net (93.55%), U-Mamba (92.46%), and SegMamba (92.76%). The improvements are statistically significant (*p <* 0.05) across all comparisons, indicating the robustness and clinical relevance of the proposed approach.

The ablation study confirms that each component—frequency branch, cross-domain attention, and adaptive router—contributes meaningfully to the overall performance, with a cumulative gain of 1.27% Dice over a strong spatial-only baseline. This validates the design choice of decomposing the problem into complementary spatial and frequency representations and fusing them adaptively based on local anatomical characteristics.

### 5.2 Thin-Wall Region Enhancement and Clinical Implications

The statistically significant improvement in thin-wall regions (*p* = 0.004) warrants careful interpretation regarding potential clinical implications. Nasal and paranasal sinus surgeries require millimeter-level precision to minimize risks such as orbital injury, vascular damage, or cerebrospinal fluid leaks. Our results suggest that frequency-based boundary enhancement may offer particular advantages in these high-risk areas, with improvement magnitude in thin-wall regions (+0.77%) exceeding overall gains (+0.52%).

#### Anatomical Context

The 41.81% prevalence of thin-wall regions in our dataset reflects the anatomical reality of sinonasal structures, where bone thickness frequently falls below 2mm near critical neurovascular structures. Traditional spatial-only networks achieve 90.57% Dice in these regions, while multi-scale frequency analysis reaches 91.34%. While this improvement appears modest in absolute terms, it may be clinically meaningful given the critical nature of these boundaries.

#### Targeted vs. Global Improvement

Notably, the frequency branch shows differential benefits: significant improvement in thin-wall regions (*p* = 0.004) but minimal change in thick-wall regions (*p* = 0.189, 99.98% vs. 99.98%). This pattern suggests that wavelet decomposition provides complementary information specifically relevant to boundary-adjacent areas, rather than uniformly enhancing all regions. We interpret this as evidence supporting our hypothesis that frequency-domain features capture structural details that spatial features alone may miss.

#### Limitations and Future Validation

We acknowledge several limitations that temper these findings:

- **Sample size:** Our test set comprises 20 cases from a single institution. Multi-center validation with larger cohorts is needed to confirm generalizability.
- **Clinical endpoints:** Our metrics (Dice, HD95) are surrogate measures. Prospective clinical studies evaluating surgical outcomes would be necessary to establish actual clinical benefit.
- **Thin-wall definition:** The 2mm threshold is chosen based on anatomical considerations but remains somewhat arbitrary. Sensitivity analysis across different thresholds (1mm, 1.5mm, 3mm) could provide additional insights.
- **Inter-observer variability:** Ground truth annotations may have inherent uncertainty in thin-wall regions. Analysis of inter-annotator agreement would strengthen interpretation.

Despite these limitations, the consistency of thin-wall improvement across 90% of test cases and the statistical robustness (*p* = 0.004) suggest that the observed effect is not merely due to chance.

### 5.3 Boundary Precision and Clinical Translation

The convergence of multiple evaluation perspectives—volumetric overlap (Dice), thin-wall regions, and surface distances—provides complementary evidence for AFS-DSN’s potential clinical utility. However, several caveats warrant discussion:

#### Multi-scale Consistency

Improvements appear consistent across volume (Dice +0.52%, *p* = 0.025), thin-wall regions (+0.77%, *p* = 0.004), and surface boundary overlap (Surface Dice @1mm +0.61%, *p* = 0.010). This pattern suggests genuine boundary enhancement rather than metric-specific artifacts. However, we note that all three metrics are computed from the same ground truth annotations, which may introduce correlated evaluation bias.

#### Surgical Relevance

The 0.626mm average surface error falls within the clinically acceptable range for endoscopic sinus surgery, while HD95 of 1.189mm suggests that worst-case deviations remain within a millimeter-scale range. However, we emphasize that these are computational metrics derived from expert annotations. The relationship between segmentation metrics and actual surgical outcomes (e.g., complication rates, revision surgery frequency) remains to be established through prospective clinical trials.

#### Precision Targeting

Statistically significant gains at 1mm tolerance (*p* = 0.010) but not 2mm tolerance (*p* = 0.265) suggest that frequency branch benefits may concentrate at fine scales where traditional spatial features plateau. This aligns with the regime most critical for avoiding surgical complications.

#### Generalizability Concerns

All metrics are evaluated on a single-institution dataset (n=20 test cases) with annotations from a single clinical team. Multi-center validation with diverse imaging protocols, scanner manufacturers, and patient populations is essential to confirm that these improvements generalize beyond our specific dataset.

In summary, while computational metrics suggest enhanced boundary precision, we emphasize that clinical utility must be validated through prospective studies evaluating surgical outcomes, surgeon confidence, and workflow integration. The current results provide preliminary evidence supporting further clinical investigation but should not be interpreted as definitive proof of clinical superiority.

### 5.4 Advantages of Frequency-Domain Processing

The success of AFS-DSN can be attributed to several factors related to frequency-domain modeling:

1. **Complementary representations:** Spatial features primarily capture local intensity patterns and textures, while frequency features capture global structural information and edge details across multiple scales. By processing both representations in parallel, AFS-DSN can leverage the strengths of each domain.
2. **Multi-scale analysis:** The use of multiple wavelet bases (db1, db2, db4) with different frequency characteristics allows the network to capture features at various scales simultaneously, which is particularly beneficial for sinonasal structures that exhibit both large-scale anatomical variations and fine-scale boundary details.
3. **Adaptive fusion:** The content-aware adaptive router enables the network to dynamically balance spatial and frequency information based on local complexity. In regions with clear boundaries and high contrast, spatial features may dominate; in regions with thin walls or subtle intensity transitions, frequency features can provide critical edge information.

### 5.5 Comparison with Transformer and Mamba Methods

Recent Transformer-based (UNETR, Swin-UNETR) and Mamba-based (U-Mamba, SegMamba) methods have achieved strong performance by capturing long-range dependencies through selfattention or state-space modeling. However, these methods still operate predominantly in the spatial domain and do not explicitly model frequency-domain information. Our results suggest that frequency-domain processing provides complementary benefits that are orthogonal to long-range dependency modeling. Future work could explore combining Transformer/Mamba architectures with frequency branches to further improve performance.

### 5.6 Parameter Efficiency and Clinical Feasibility

Although AFS-DSN introduces substantially more parameters (414.57M) than traditional CNN-based segmentation models and is larger than Transformer-based methods such as UNETR (92.8M), this increase is mainly attributable to the frequency branch, which processes multiple wavelet subbands in parallel to capture boundary-relevant features at different scales. Importantly, the added model capacity yields measurable improvements in clinically critical boundary-focused evaluations, including thin-wall Dice (+0.77%, *p* = 0.004) and Surface Dice at 1mm tolerance (+0.61%, *p* = 0.010), rather than global volumetric overlap alone (+0.52% Dice).

To directly address the performance-versus-complexity trade-off, we introduce AFS-DSN-Lite, a parameter-efficient variant that replaces the high-capacity frequency fusion block with depthwise separable convolutions. AFS-DSN-Lite achieves 94.37% Dice with only 27.41M parameters—a 93.5% reduction from the full model—while requiring 488.05 GFLOPs and 260.0 ms inference time. Notably, AFS-DSN-Lite marginally outperforms the full model in overall Dice (94.37% vs. 94.34%), suggesting that the depthwise separable design provides sufficient capacity to capture the relevant frequency features for this task. This result demonstrates that the performance gains of frequency-domain processing are not dependent on the large parameter count of the full fusion block.

### 5.7 Limitations and Future Work

#### Binary vs. multi-class formulation

In this work, all five sinonasal structures are merged into a single foreground class, resulting in a binary segmentation formulation. This design choice is motivated by clinical requirements for surgical planning and navigation, where accurate delineation of the overall nasal–sinus complex boundary is prioritized over per-structure volumetry. However, we acknowledge that this formulation limits direct analysis of individual anatomical structures and may not be suitable for applications requiring structure-specific measurements. Extending AFS-DSN to a multi-class segmentation framework is an important direction for future work.

Several limitations should be acknowledged:

1. **Dataset size:** The NasalSeg dataset contains 130 cases, which is relatively small for deep learning standards. Although our model generalizes well to the independent test set, evaluation on larger and more diverse datasets would strengthen the findings. To address the concern of single-split evaluation, we conducted 3-fold cross-validation on the NasalSeg dataset. AFS-DSN achieved a mean Dice of 94.59% ± 0.31% across the three folds (Fold 1: 94.25%, Fold 2: 94.50%, Fold 3: 95.01%), demonstrating that the reported performance is not the result of a favorable data split but reflects consistent generalization capability.
2. **Anatomical scope:** The current study focuses on five major sinonasal structures. Future work could extend the framework to segment additional fine-grained structures (e.g., individual turbinates, ostia, septum) for more detailed surgical planning.
3. **Computational cost:** While inference time is acceptable, training time (12 hours for four stages) could be reduced through more efficient training strategies or by leveraging pretrained models from related tasks.
4. **External validation:** Validation on external datasets from different institutions with varying imaging protocols would further demonstrate the generalizability and robustness of AFSDSN.

Future research directions include:

- Combining frequency-domain processing with Transformer or Mamba architectures for enhanced long-range modeling
- Extending AFS-DSN to multi-organ segmentation tasks in head-and-neck imaging
- Incorporating uncertainty quantification to identify challenging cases requiring manual review
- Developing lightweight variants for real-time or resource-constrained clinical deployment

## 6 Conclusion

This paper proposes AFS-DSN, an Adaptive Frequency–Spatial Dual-Stream Network for automatic segmentation of the nasal cavity and paranasal sinuses from 3D CT volumes. The key innovation lies in explicitly modeling and adaptively fusing spatial and frequency-domain representations through multi-scale wavelet decomposition, cross-domain attention, and content-aware adaptive routing. Extensive experiments on the NasalSeg dataset demonstrate that AFS-DSN achieves promising performance, with a Dice score of 94.34% v 2.30% and HD95 of 1.189 mm on the independent test set, significantly outperforming strong baselines including nnU-Net, UNETR, Swin-UNETR, U-Mamba, and SegMamba. We further introduce AFS-DSN-Lite, a parameter-efficient variant achieving 94.37% Dice with only 27.41M parameters, and validate robustness through 3-fold cross-validation (mean Dice: 94.59% ± 0.31%). Ablation studies confirm that the frequency branch, cross-domain attention, and adaptive router each contribute meaningfully to the overall performance, while generalization analysis demonstrates robust performance across training, validation, and test splits.

The success of AFS-DSN suggests that explicitly modeling frequency-domain information and adaptively fusing it with spatial features is a promising direction for medical image segmentation, particularly for tasks involving complex anatomical structures, thin boundaries, and low-contrast regions. Future work will focus on three key directions: (1) developing lightweight variants through parameter-efficient fusion strategies to facilitate real-time clinical deployment; (2) conducting external validation on multi-institutional datasets to confirm robustness across diverse imaging protocols; and (3) investigating hybrid architectures that synergize our frequency-domain analysis with long-range modeling mechanisms (e.g., Transformers) to further enhance structural consistency in complex anatomical topologies.

## Data Availability

The CT imaging data used in this study were obtained from the publicly available NasalSeg dataset, an open-access nasal cavity and paranasal sinus CT dataset with voxel-wise annotations of five anatomical structures (https://zenodo.org/records/13893419). Due to licensing and privacy considerations, we do not redistribute the original images. Processed data and trained model weights generated in this work are available from the corresponding author upon reasonable request.

## Supplementary Material

### Wavelet Scale Robustness

### Parameter Breakdown Analysis

These results confirm the incremental benefits of multi-scale decomposition and the robustness of the full AFS-DSN architecture.

## Acknowledgment

This study was partially supported by the National Science and Technology Council (NSTC) undergraduate student research projects under Grant NSTC-114-2813-C-182-053-E and NSTC-113-2221-E-182-022. The authors also gratefully acknowledge the creators of the NasalSeg dataset for providing open-access nasal cavity and paranasal sinus CT data that enabled the experimental evaluation in this paper.

